# Bionnica: A Deep Neural Network Architecture for Colorectal Polyps’ Premalignancy Risk Evaluation

**DOI:** 10.1101/2024.06.19.24309153

**Authors:** Diogen Babuc, Todor Ivaşcu, Melania Ardelean, Darian Onchiş

## Abstract

The third most prevalent cancer nowadays is colorectal cancer. Colonoscopy is an important procedure in the stage of detection of polyps’ malignancy because it helps in early identification and establishes effective therapy. This paper explores specific deep-learning architectures for the binary classification of colorectal polyps and considers the evaluation of their premalignancy risk. The main scope is to create a custom-based deep learning architecture that classifies adenomatous, hyperplastic, and serrated polyps’ samples into benign and premalignant based on images from the colonoscopic dataset. Each image’s output is modified through masked autoencoders which enhance the classification performance of the proposed model, called *Bionnica*. From the four evaluated state-of-the-art deep learning models (ZF NET, VGG-16, AlexNet, and ResNet-50), our experiments showed that ResNet-50 and ZF NET are most accurate (above 84%), with ResNet-50 excelling at indicating patients with premalignant colorectal polyps (above 92%). ZF NET is the fastest at handling 700 images. Our proposed deep learning model, *Bionnica*, is more performant than ZF NET and provides an efficient classification of colorectal polyps given its simple structure. The advantage of our model comes from the custom enhancement interpretability with a rule-based layer that guides the learning process and supports medical personnel in their decisions.

## 1 Introduction

The third most frequent cancer malignancy and the second most deadly type of cancer is colorectal cancer [1], albeit its exact etiology is not always known [2]. However, there are several risk factors linked to a higher chance of acquiring colorectal cancer [3], including age (people over 50 are at a higher likelihood of developing colorectal cancer than those under 50), family history (certain inherited disorders or a history of colorectal cancer in the family can raise the risk), personal history (having had colorectal cancer in the past or having specific types of polyps can make it that an individual will get it again), Crohn’s disease and ulcerative colitis (inflammatory bowel illnesses), and lifestyle factors (diets, sedentary lifestyle, obesity, smoking, excessive alcohol consumption) [4].

Regular screening is crucial for the early detection of colorectal cancer and colonoscopy is viewed as the epitome for screening; a long flexible fiber-optic tube (with a camera at one end) is inserted through the anus that allows visualization of the mucous lining, to detect polyps and colorectal cancer [5]. A colonoscopy is a highly recommended screening tool, especially for people with risk factors or people over a certain age. Early discovery is essential for effective treatment.

Automated systems can process a large number of images quickly, allowing for a more efficient evaluation of colorectal polyps. This is particularly beneficial in screening programs with a large volume of data. The identification of premalignant polyps can replace a biopsy up to a certain point. In the current study, convolutional neural networks (CNN) such as the Zeiler-Fergus neural network (ZF NET) [6], Visual Geometry Group 16 (VGG-16) [7], AlexNet [8], and residual neural network (ResNet-50) [9] were used, as they have consistently demonstrated superior performance in previous research focused on evaluating the risk of premalignancy of colorectal polyps. This choice is based on the extensive body of specialized literature, where these deep CNN models have consistently shown remarkable efficacy. We created a custom convolutional network that, even though it has few layers, functions well, being fast in training and, if necessary, retraining. By adding a rule-based layer, we incorporate medical knowledge to increase accuracy, precision, sensitivity, and ROC-AUC. The main contribution of this paper is creating an adjusted deep neural network that can help in detecting the risk of malignancy of colorectal polyps and be more performant than other state-of-the-art deep learning models in terms of evaluation metrics and execution time.

The main objectives of this paper are:

1. Understanding state-of-the-art deep learning architecture and important features for bioimage processing,
2. Building a custom-based deep learning architecture, *Bionnica*, for classifying colorectal polyps into premalignant and benign.
3. Evaluation of all deep learning approaches considering performance metrics, which further estimates the colorectal polyps’ premalignancy risk, and execution time for architectures’ construction and data processing, and calculating results.

This paper is structured as follows. In Section 2, we discuss colonoscopy screening and feature selection. In Subsection 2.1, we elaborate on the polyps analysis and give some concrete examples of malignant polyps. We analyze the features obtained during the virtual biopsy. In Subsection 2.2, we go into the deep learning approaches, where we examine the modern architecture that can significantly improve the detection factor of polyps. We briefly define them and indicate some related works. In Section 3, we elaborate on our deep learning architecture. In Subsection 3.1, we explain how to manipulate data for the preprocessing and processing parts of the experiment. In Subsection 3.2, we give details on the concrete implementation logistics of the deep model. We confer on our authentic model, which tries to surpass existing deep architectures for the risk of malignancy of colorectal polyps. In Section 4, we discuss the graphical and numerical results and compare them. Afterward, we deliberate on the evaluation metrics and explain the importance of the proper selection of deep architectures for the digital biopsy using automated colonoscopy. We appraise the best-performant models in terms of execution time and performance evaluation metrics. In Section 5, we create a recap of the paper, indicating the advantages of our research, but also specifying the limitations, and giving the potential solutions for them.

## 2 Background and Related Works

Important features of bioimage processing in colonoscopy include advanced image analysis algorithms that aid in the colorectal polyps’ risk of malignancy, lesions, and abnormalities, as well as real-time image enhancement techniques that improve the visualization of the colon’s mucosa, contributing to more accurate diagnoses and early intervention in colorectal diseases [10]. Additionally, automated computer-aided diagnosis systems are becoming increasingly essential for assisting healthcare providers in interpreting colonoscopy images, and streamlining the detection and characterization of colorectal lesions [11]. Computer-aided systems can analyze medical images with a high degree of accuracy and consistency. They are not prone to human errors or variations in interpretation, which can be crucial in identifying subtle features that may indicate malignancy [12].

### 2.1 Important Features in Colonoscopy

Colorectal lesions refer to abnormal growths or changes in the colon and rectum. These lesions can be non-cancerous or cancerous. Lesions are typically identified through various medical imaging techniques, such as colonoscopy, sigmoidoscopy, barium enema, or computed tomography scans [10]. Polyps are small, fleshy growths on the inner lining of the colon or rectum. These are usually removed during a colonoscopy to prevent the risk of cancer. Conditions like Crohn’s disease and ulcerative colitis can cause chronic inflammation in the colon and rectum, leading to the development of lesions that can be pre-cancerous [4], leading to the development of dysplastic lesions that represent precursor lesions of colorectal cancer.

Adenomas are a particular kind of polyp that can develop into carcinoma. The colon and rectum are the places where these precancerous growths are most frequently seen. Adenomas can range in size and appearance, and according to their form, they can be categorized as tubular, villous, or tubulovillous adenomas [2]. The size, number, morphology, and other factors of polyps can influence their development into colorectal cancers. This is why one of the most effective methods for preventing colorectal cancer is the excision of adenomas during a colonoscopy [13]. Usually, hyperplastic polyps are not thought to be malignant [14]. Some hyperplastic polyps (Fig. 1), nevertheless, can contain serrated features, which could enhance their likelihood of developing cancer [2]. To prevent them from turning into cancer, large or serrated hyperplastic polyps are more likely to be observed or removed during a colonoscopy. Certain adenomas and hyperplastic polyps may exhibit serrated characteristics, which implies that, under the microscope, they may seem sawtoothed or serrated [15]. Serrated polyps (Fig. 2) are particularly intriguing since some of them are more prone to develop into cancer. In fact, serrated polyps are linked to a subtype of colorectal cancer called *serrated neoplasia pathway* [13]. To decrease the probability that these routes would eventually lead to cancer, it is crucial to spot and keep an eye on serrated polyps during colonoscopy.

**Fig. 1.**
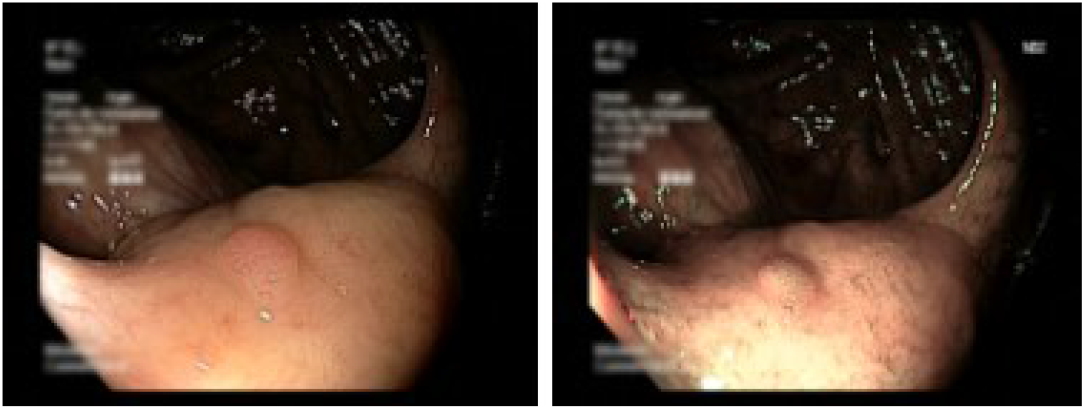
Hyperplastic polyp 08 from the colonoscopic dataset [16] captured with a white light frame and narrow band imaging, respectively.

**Fig. 2.**
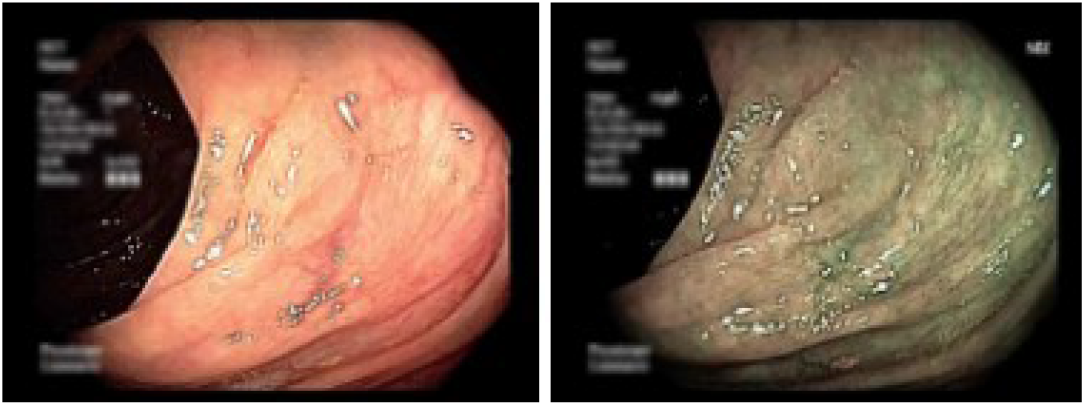
Serrated polyp 07 from the colonoscopic dataset [16] captured with a white light frame and narrow band imaging, respectively.

During a colonoscopy, a white light frame is essentially the standard mode of visualization used by the endoscope [10, 17]. The white light frame in a colonoscopy provides a clear, well-lit view of the inside of the colon and rectum. This white light is used to illuminate the mucosal lining of the colon, making it easier for the healthcare provider to examine the area for any abnormalities, such as polyps, lesions, or signs of inflammation [10]. It permits the endoscopist to carefully inspect the entire length of the colon and rectum for any signs of disease, including colorectal polyps. The white light frame provides real-time imaging of the colon’s interior on a monitor [18]. If any suspicious areas or lesions are identified during the colonoscopy, the endoscopist can use specialized instruments to take biopsies or remove polyps. This is typically done under white light guidance [19].

Narrow Band Imaging (NBI) is based on the principle that different wavelengths of light are absorbed by tissues to varying degrees. It uses narrow-bandwidth light in two specific wavelengths (415 nm and 540 nm) to enhance the contrast between blood vessels and the surrounding mucosa [19]. This allows for a more detailed inspection of the mucosal pattern and the blood vessels in the gastrointestinal tract [20]. NBI can improve the detection rate of various conditions, including precancerous polyps, early-stage cancers, and other gastrointestinal abnormalities. It is particularly valuable in the detection and characterization of lesions in the esophagus, stomach, and colon [21].

Camera calibration is a crucial process in computer vision, computer graphics, and various other fields where cameras are used for image and video processing [10]. It involves determining the mathematical relationship between the 3D world and the 2D image plane, enabling accurate measurement and analysis of objects and scenes [22]. Camera calibration allows for the conversion of pixel measurements in images to real-world measurements in physical units. This is essential for tasks like object size estimation, distance measurement, and 3D reconstruction. Cameras may exhibit lens distortions, such as radial distortion and tangential distortion. Calibration provides correction parameters that can be applied to images to remove these distortions, resulting in more accurate and distortion-free images [23].

### 2.2 Deep Learning Approaches

Over the years, several CNN architectures have been developed, such as segmentation neural network [24], AlexNet, ZF NET, VGG-16, GoogLeNet [25], and ResNet-50. Despite the various CNN architectures, the models share common components, such as the pooling layer, the convolutional layer, and the fully connected layer. To maintain information, fully connected networks use a skip connection, a sampling path, and downsampling connections. Deep learning and region proposal networks are combined in faster *Region-based Convolutional Neural Networks*, making them useful for the precise and effective classification of polyps [4].

#### Residual Neural Network (ResNet-50)

ResNet-50 is typically made up of many residual blocks stacked on top of each other [26]. The number of residual blocks and their depth can be adjusted based on the specific task and dataset. ResNet-50 architectures are widely used in image classification, object detection, face recognition, and other computer vision tasks. ResNet-50 has achieved state-of-the-art outputs on colonoscopy image classification and object detection tasks [28]. ResNet-50’s skip connections make it easier to train very deep networks without suffering from vanishing gradients. This allows the construction of much deeper neural networks than previously possible [29].

ResNet-50 achieved state-of-the-art performance on various image classification benchmarks [9]. Its deep architecture with residual connections allows it to learn more discriminative features and capture complex patterns in images, leading to higher accuracy compared to shallower architectures. Despite its depth, ResNet-50 can be trained efficiently with reasonable memory usage. The skip connections in ResNet-50 help alleviate the vanishing gradient problem, allowing for more stable and efficient training compared to architectures without such connections. While ResNet-50 addresses the vanishing gradient problem and enables the training of very deep networks, it comes at the cost of increased computational complexity.

Deeper architectures require more computation during both training and inference, which can be computationally expensive, especially for resource-constrained environments [29]. These could be some limitations of the architecture.

The authors of [27] gained a 95.79% F1-score and 91.62% Jaccard Index (Table 1), while the authors of [30] received the F1-score of 91.57%, an accuracy of 98.82%, a sensitivity of 98.28%, and a specificity of 98.68% (Table 2), from a dataset [31] known as *The Cancer Imaging Archive* in the context of *CT Colonography* [32].

**Table 1.** The performance evaluation metrics, obtained by authors from [27], applied on the *CVC-ClinicDB*.

| % | Res-U-Net |
| --- | --- |
| **F1-score** | 95.79 |
| **Jaccard Index** | 91.62 |

**Table 2.** The performance evaluation metrics obtained by authors from. [30] **for ResNet-50 and applied on CT images.**

| % | ResNet-50 |
| --- | --- |
| **F1-Score** | 91.57 |
| **Accuracy** | 98.82 |
| **Sensitivity** | 98.28 |
| **Specificity** | 98.68 |

#### AlexNet Architecture

The AlexNet deep learning model has various important uses right now, including the risk estimation of colorectal polyps and other medical images. AlexNet’s deep architecture with multiple convolutional and pooling layers enables it to learn hierarchical features from images [8], leading to high accuracy in classification tasks. AlexNet popularized ReLU as activation functions instead of traditional sigmoid or *tanh* functions [33]. ReLU activation functions accelerate the training process by mitigating the vanishing gradient problem and enabling faster convergence compared to saturating nonlinearities. The deep and complex architecture of AlexNet may reduce model interpretability, making it challenging to understand how the network makes predictions or which features are essential for classification [30]. Understanding the internal representations learned by the network can be more challenging in architectures with many layers and parameters. Using the following dataset [34], the authors of [30] got a 44% precision and 63.78% sensitivity, while for the work [33], this model obtained the F1-score value bigger than 80%, precision of 88.3%, and a sensitivity of 74.8%. This architecture is excellent in classifying images using hierarchical class information [35]. They can group images into precise subcategories, which improves diagnostic precision. This architecture is used for object detection [36], particularly for the location of polyps in medical images [37], but also for bioimage classification. AlexNet architecture is capable of tracking objects across time while preserving the target’s consistent appearance. This helps locate polyp regions among a series of pictures, improving colonoscopy diagnostic performance [38]. Additionally, they carry out operations such as super-resolution, compressive sensing, and image denoising to improve the quality of the images and the ability to discriminate [42].

#### Visual Geometry Group 16 (VGG-16)

VGG-16 is specifically named because it consists of 16 weight layers. VGG-16 is characterized by its deep structure with small 3 x 3 convolutional filters. VGG-16 has a straightforward and uniform architecture, making it easy to understand and implement [38].

Considering transfer learning, pre-trained VGG-16 models, trained on large-scale datasets such as *ImageNet*, are readily available. These pre-trained models can be finetuned on smaller datasets or adapted to different tasks with transfer learning, where the learned features from the ImageNet dataset can be leveraged for various image-related tasks, such as object detection, segmentation, and localization [7]. VGG-16 provides good generalization performance across different datasets and domains. Its ability to learn hierarchical features allows it to capture and generalize patterns from diverse images, making it suitable for a wide range of image classification tasks without extensive architecture modifications [38, 39].

The authors of [26] obtained a precision of 73.6% and sensitivity of 86.3% from the CVC colon datasets. Also for these datasets, in another study [40], the authors got the F1-score of 98.3% and intersection-over-union of 96.95%, which are performant results.

#### Zeiler-Fergus Neural Network (ZF NET)

ZF NET is a deep learning architecture, most similar with the proposed custom-based model. For ZF NET we also introduce and analyze the formulas for some layers because it is the most performant deep learning architecture from all of the state-of-the-art models. ZF NET consists of five convolutional layers, followed by three fully connected layers. The network uses rectified linear units (ReLU) as activation functions and employs techniques like dropout and local response normalization to improve generalization performance.

ZF NET achieved state-of-the-art performance in the ILSVRC 2013 competition, surpassing previous architectures in terms of accuracy [6]. One notable aspect of ZF NET is that it provided insights into the features learned by the convolutional layers through visualizations of the learned filters. The visualization techniques used in ZF NET helped researchers better understand how deep learning models perceive and interpret images. In contrary, ZF NET still requires significant computational resources for training and inference. This computational complexity could be a disadvantage, particularly for applications with limited computational resources. Like many deep learning models, ZF NET is susceptible to overfitting, especially when trained on small datasets or datasets with limited diversity. Overfitting can lead to reduced generalization performance on unseen data, which undermines the model’s effectiveness in real-world applications.

In the preliminary experiment of the study [37], 10% of the database [41] was set aside to evaluate the efficiency of various networks. In terms of sensitivity, the pre-trained ZF NET beat the other networks in general. To improve resilience and prevent overfitting, the network included techniques such as Max-Pooling, ReLU activation, and dropout. The final fully connected layer was modified to produce two classes: *Polyps* and *No Polyps* [42]. The convolutional layer in ZF NET performs convolutions between input images and learnable filters. Let’s denote the input tensor as *X* and the filter tensor as *W* (see (1)). Variables *i* and *j* are the indices of the spatial dimensions of the input feature map, *X*, while *m* and *n* are the indices of the spatial dimensions of the filter, *W*. Variables *m* and *n* represent the position of the element in the 2D filter. The convolution operation at a particular spatial location can be expressed as [43]:

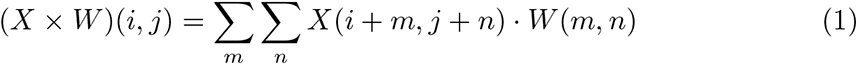

This operation involves sliding the filter over the input tensor, computing element-wise products, and summing them up. ZF NET uses max pooling, which involves downsampling the spatial dimensions of the input tensor by selecting the maximum value from each local region (see (2)). If we denote the pooling window size as *s*, and *p* and *q* are the indices used for pooling window positions, the pooling operation can be expressed as:

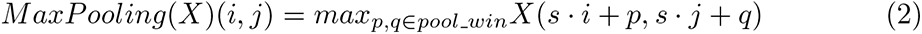

where *p, q pool win* indicates that *p* and *q* take values from the set of indices defined by the pooling window. In the fully connected layers, matrix multiplication is performed between the input vector and the weight matrix, followed by the addition of biases. Let *X* be the input vector, *W* the weight matrix, and *b* the bias vector (see 3)). The fully connected operation is:

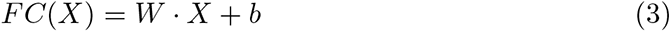

These operations are foundational in understanding the linear algebra involved in the forward pass of ZF NET. The network typically ends with softmax activation for classification tasks. Considering layer-by-layer stage processing, a sequential deep model is initialized. This model adds a series of convolutional layers with different filter sizes, activation functions (ReLU), and pooling layers. After convolutional layers, the model is flattened, and fully connected layers are added. The flattened layer reshapes the 3D tensor into a 1D tensor, preparing it for the fully connected layers (Fig. 3). This can be seen as the flattening of a matrix into a vector. The categorical cross-entropy loss function and Adam optimizer are used to compile the model. Accuracy is used as a metric [44]. A CNN study, based on ZF NET deep learning architecture [45] attained a high ROC-AUC, 0.991, with an accuracy of 96.4%. It has a 7% false positive rate. Another study [46] contrasted endoscopic visual assessment with the use of a CNN (the structure of ZF NET). Both methods demonstrated comparable precision (87.3% vs. 86.4%), but the second study had a greater sensitivity value (87.6% vs. 77%) and an accuracy of (85.9% vs. 74.3%).

**Fig. 3.**
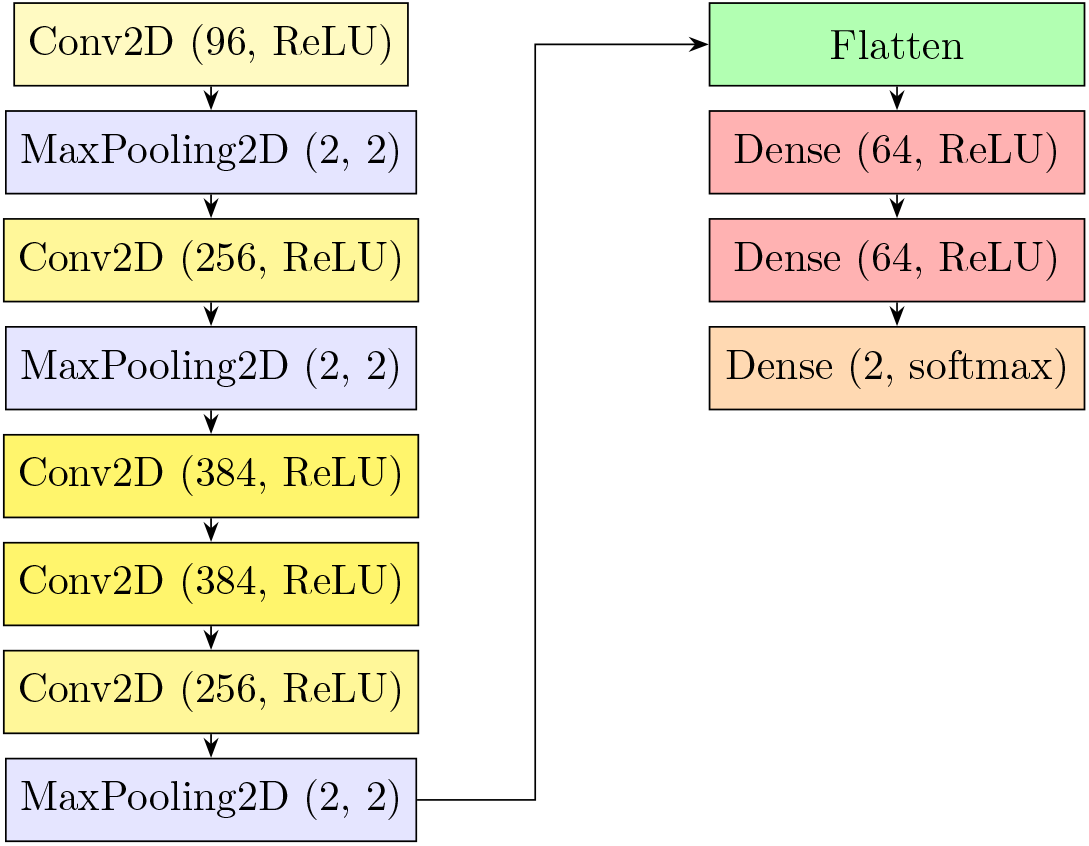
ZF NET deep learning architecture for classifying colorectal polyps into premalignant and benign.

All analyzed state-of-the-art algorithms use deep learning architectures. Sequential models with convolutional layers are initialized and established. Each layer has a ReLU or softmax activation function, and from 32 to 384 filters. Each model has at least one max-pooling layer added to the convolutional one to reduce logically the number of features and observations. Also, fully connected layers are integrated into the models, with a dropout function to prevent overfitting. The output layer in general has 64 neurons for the classification problem with the softmax activation function.

### 2.3 Masked Autoencoders

Masked autoencoder (MAE) is a method used for supervised or unsupervised learning. It is particularly useful for image reconstruction and feature extraction. It works by masking parts of the input and, afterwards, training the model to reconstruct the missing parts [47]. This is beneficial for the model that learns features and patterns within the data. A MAE can be used to analyze colorectal polyps from colonoscopic images, to assess the risk of premalignancy. The process of implementing this method contains data preparations, model architecture, training part, feature extraction, risk assessment, and model’s validation and testing.

Images are usually normalized to ensure the consistency. Parts of the input images can be either randomly or systematically masked. The model learn to reconstruct these masked regions. This will lead to a better understanding of the contextual information of the image.

This principle is trained using gradient descent and backpropagation to minimize the reconstruction loss. It is composed of an encoder and a decoder. So,it compresses the input into a compact, latent representation. Also, it reconstructs the original image from the latent representation, to fill the masked portions [48]. Considering the supervised training, the classifier is trained on labeled data where the premalignancy status of polyps is known. Techniques such as k-fold cross-validation can be used to ensure the model’s robustness.

Challenges of this approach are the following. MAE cannot interpret the feature in a clinically meaningful way. Also, MAE requires significant computational resources for training, especially when there are high-resolution medical images, indicating that a solution could be achieved by downscaling the images.

## 3 Proposed Approach

The initial phase of the implementation primarily centers on data preprocessing, data segmentation, feature selection, dataset splitting, and the categorization and classification of colorectal polyps. By employing threshold values, patients are categorized into those at risk and those not at risk of developing colorectal cancer (having a premalignant polyp or not).

### 3.1 Dataset Analysis and Data Preprocessing

The dataset used for this paper is the publicly available dataset on the *Depeca* platform [16]. The examination of the dataset’s features is essential as they indicate the focus and significance of the study. It consists of multiple captures of different polyps types records. To examine the dataset, we listed the columns obtained from the bioimage processing in the colonoscopic dataset. In total, there are 700 images related to serrated, adenomatous, and hyperplastic polyps. A virtual biopsy framework has been developed for classifying gastrointestinal lesions without altering existing medical protocols or hardware requirements, relying solely on the availability of NBI lighting, a standard feature in colonoscopies. The system generates 3D shape reconstructions of the lesions, aiding doctors in diagnosis, to how 3D visual information is utilized in other medical research domains. Moreover, a colonoscopic video database has been curated and published, offering interdisciplinary research collaboration between computer scientists, biomedical engineers, and medical professionals. This dataset includes colonoscopic videos captured under both white light and NBI modes, accompanied by ground truth annotations derived from endoscopist image inspection. Also, histological analysis is implicated, as well as camera calibration information [10]. To address class imbalance in datasets, one commonly employed method is through resampling techniques, namely oversampling and undersampling. Oversampling involves replicating instances from the minority class, while undersampling involves reducing instances from the majority class until a balanced distribution is achieved. However, it is imperative to rigorously assess the implications of these techniques on model performance, and validation through cross-validation or other methods is essential to ensure the effectiveness of the chosen approach.

Considering the image acquisition, masked autoencoders obtained specifically-formatted images of colorectal polyps [49] that help model to achieve performant results (see Fig. 4). The annotation was also made as a part of data preprocessing, by labeling the images with information about the presence and type of polyps, and their malignancy status. The images are normalized for consistent contrast and brightness [50]. Analysing the masking strategy, the mask is applied in a structured pattern. For the images, an encoder is implemented to compress the input images into a latent representation, and a decoder to reconstruct the masked portions. Using the latent representation as input features for the classification model, the premalignancy risk of the polyp is established and categorized properly [51].

**Fig. 4.**
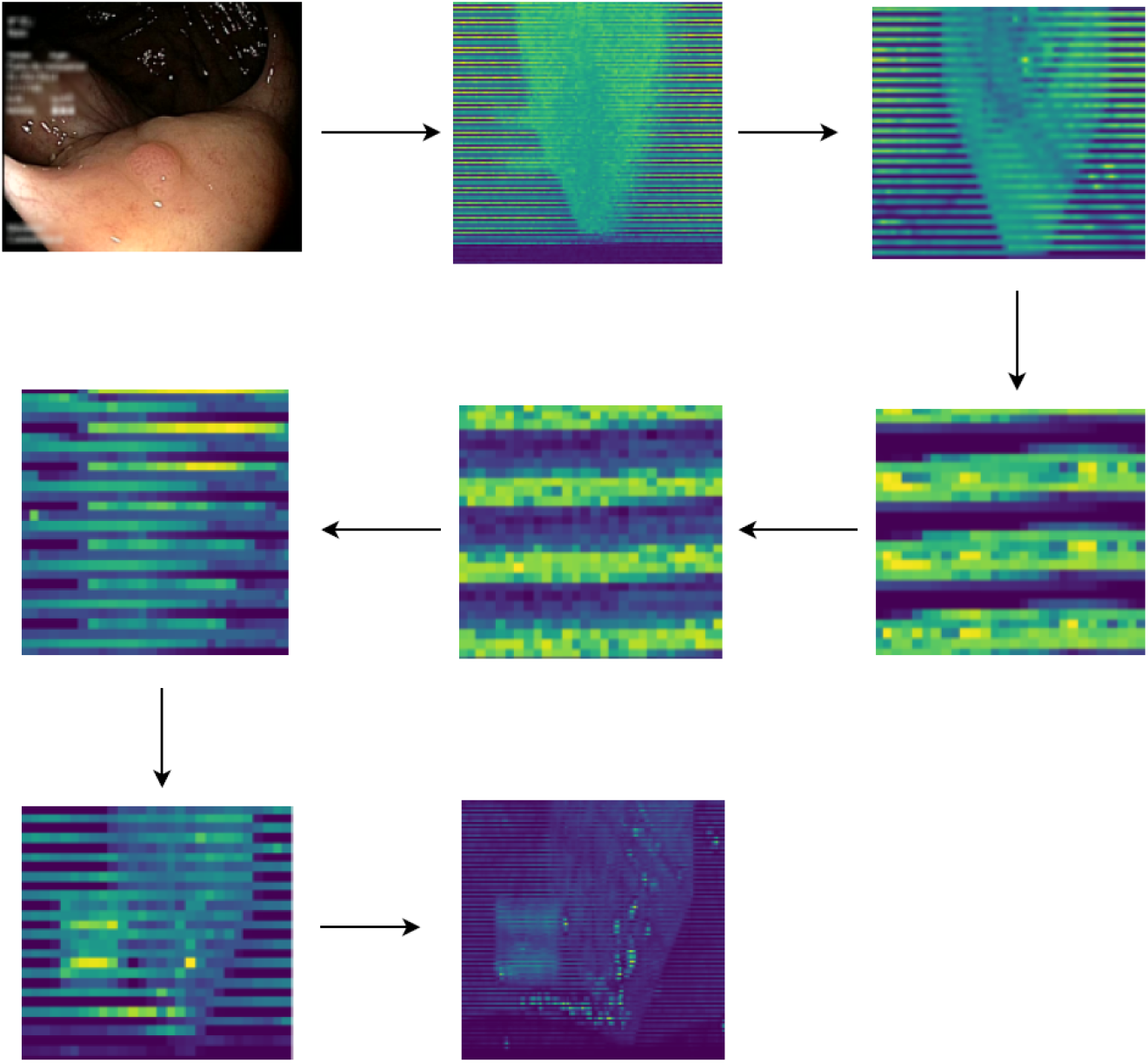
Masked autoencoders on image processing for a potential image classification’s results enhancement.

To classify individuals in terms of polyp status, cut-off values are determined for the most critical parameter [52]. This cutoff helps categorize individuals into those with premalignant polyps and those with benign polyps. The target variable uses binary classification, using 1 and 0 to signify the presence or absence of premalignant polyps following virtual colonoscopy scans. After assessing the importance of the feature set, it was decided that all columns of the dataset (excluding the target column) should be included in the independent variable. Notably, a strong correlation (above 0.7) exists between all features and the dependent variable, using point-biserial correlation which measures the correlation between a continuous variable and a binary variable.

Youden’s J Index, is a useful measure for determining an optimal cutoff value in binary classification problems [53]. It considers both sensitivity and specificity of a diagnostic test to find the threshold that maximizes the difference between true positive and false positive rates. Thus, the cutoff value, determined using Youden’s J index method, serves as an indicator of similarity between expected and model-derived values. It offers a balanced single metric that optimally combines sensitivity and specificity, aiding in the selection of an optimal cutoff value (10% in our case). To find the optimal cutoff value using Youden’s J index, it is necessary to calculate sensitivity and specificity for each possible cutoff value; then, to compute Youden’s J index for each cutoff value, using the formula: *sensitivity* + *specificity* 1, and afterward select the cutoff value that maximizes Youden’s J index [54].

The confusion matrix is important for evaluating the performance of a classifier, such as when classifying colorectal polyps into premalignant and benign. It provides a detailed breakdown of the classifier’s predictions compared to the actual ground truth labels. This breakdown allows for a better understanding of the classifier’s strengths and weaknesses, particularly in the context of medical diagnoses where accurate classification is critical [55]. A false negative is particularly concerning because it means the classifier missed identifying a premalignant polyp, potentially leading to a missed diagnosis and delayed treatment, which could be critical for the patient’s health outcome. A false positive could lead to unnecessary anxiety and additional testing or procedures for patients who do not have premalignant polyps.

True positive (TP) and true negative (TN) values indicate correct predictions, with TP being crucial for identifying actual premalignant cases and TN indicating correct identification of benign cases.

### 3.2 Construction of the Custom-Based Architecture

The proposed model is called *Bionnica*, where *Bio* comes from the biomedical decisions that this system owes. The other part of this architecture’s name comes from *neural network innovative customization with medical assistance*.

For our model, three convolutional layers are added (they can enhance the model’s capacity for hierarchical feature extraction, enabling the capture of intricate patterns and representations in the input data), which perform operations with 32, 64, and 128 filters.

Each convolutional layer captures different levels of abstraction and complexity in the input data. The first layer might learn simple patterns, such as edges and textures, while subsequent layers can learn increasingly complex features that are combinations of lower-level features. The activation function for all of them is ReLU, which helps mitigate the vanishing gradient problem, where gradients become very small during backpropagation. Max-pooling layers follow the convolution layers to reduce the spatial dimensions of feature maps, thereby reducing the complexity of the problem by capturing important features.

Here we integrate the user and item embeddings as a collaborative filtering [56, 57] technique. In a medical context, we define a hypothetical interaction scenario where users are medical practitioners and patients. The items in this scenario are colorectal images. The interactions between users and items include diagnosis decisions, image quality ratings, and any form of user engagement with the images, such as annotations. We create a set of hypothetical users by categorizing them based on different criteria, such as their experience level and medical specialty. Next, we use the available annotations on the colorectal images to define the items. These labels include the type of polyp and its severity. For example, senior doctors might have higher confidence in diagnosing images and thus provide accurate ratings and detailed comments. We also simulate different engagement levels, where some users interact with a larger number of images, providing comprehensive data. We create encodings for users and items to facilitate the collaborative filtering process. We start by assigning unique numerical values to each user and item using label encoding techniques. This encoding allows us to convert the user and item information into a format that can be used as input for embedding layers in our model. These embeddings will capture latent factors representing user preferences and item characteristics, enabling the collaborative filtering process to provide personalized classifications.

The next layer allows the output to be reshaped into a 1D vector, transitioning from the convolutional layer to a fully connected layer. We added two fully connected layers to the model [44]. The first one has 128 units (enhance model complexity and feature abstraction), with the ReLU activation function. The second one has 2 units (output units) with the softmax activation function and is added after the dropout regularisation technique (as in Fig. 5). For each polyp’s capture, a *premalignant* or *benign* value is added to a list for a single patient.

**Fig. 5.**
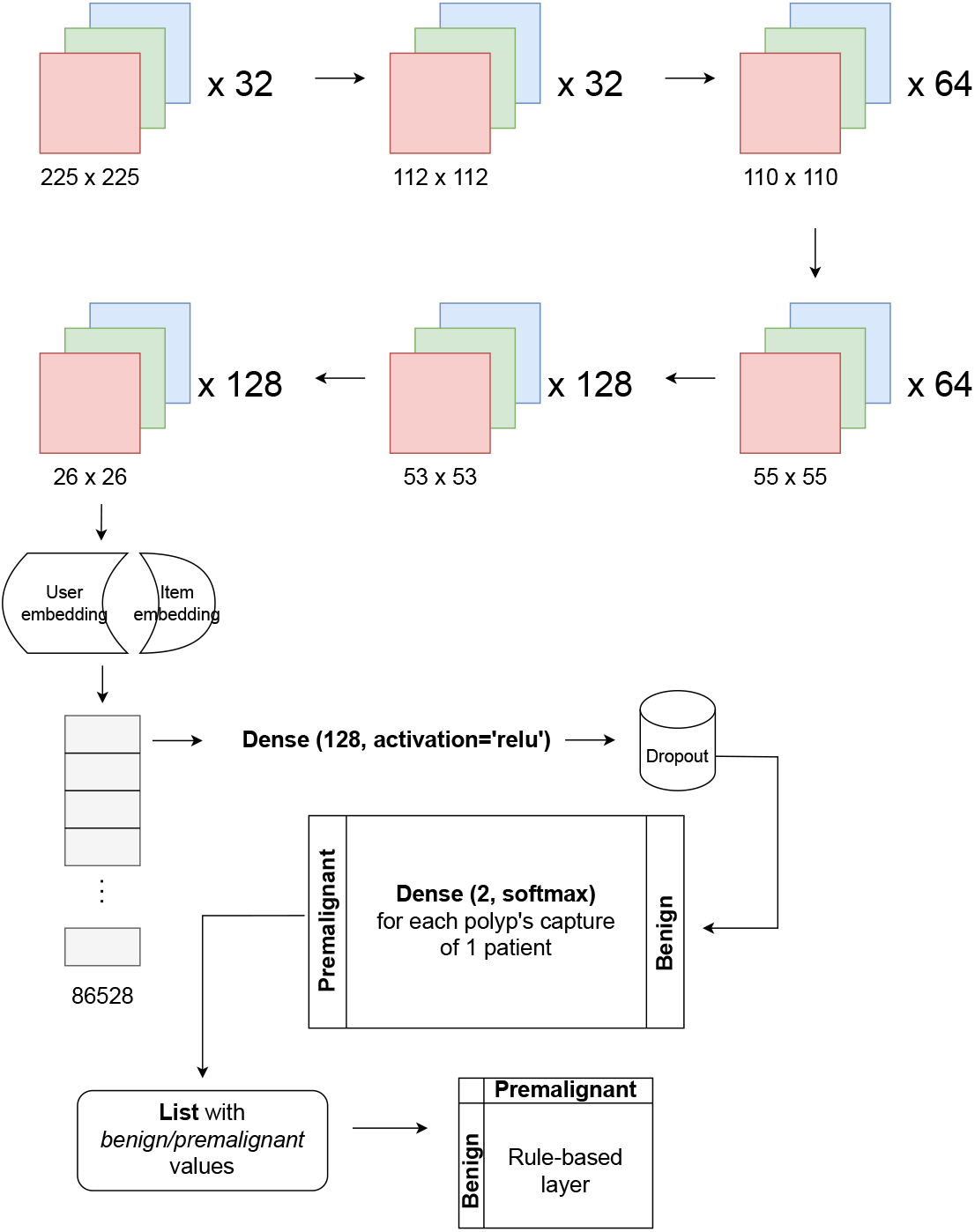
Bionnica layers and functionality illustration.

After that, a rule-based layer is added to improve the model’s performance through medical expertise and rules. For each *premalignant* value from the list, it appends 1 to the *polyp risk* target list; otherwise, it appends 0. Based on the counts of 1 and 0 (*c*1 and *c*0), it makes a decision. If value 1 appears in more than 10% of cases (obtained by Youden’s J index), it returns 1, specifying that a certain patient suffers from a premalignant polyp. Otherwise, it returns 0, indicating there is no polyp’s risk of malignancy. By specifying the percentage as a threshold, we generalize the usage of the proposed architecture. For compiling the model, we used the *Adam* optimizer (adaptive learning rates, fast convergence), specifying the learning rate of 0.001 (stable convergence, small weight updates) and used, as a loss function, the binary cross-entropy, which is suitable for the binary classification and yields good performance metrics.

Rule-based layers can enhance the interpretability of the model. By explicitly defining rules with values of 0 and 1, it becomes easier to understand how the model makes decisions. By indicating rules based on expert insights, the model can use this information to improve performance. Also, values of 0 and 1 in rule-based layers facilitate model explainability. By examining which rules are activated or deactivated for a given input, stakeholders can gain insights into the model’s decision-making process.

However, rule-based layers may not be able to capture complex relationships present in the data because rule-based layers are constrained by predefined rules. Rule-based layers are susceptible to overfitting, especially when the rules are specific or tailored to the training data. In such cases, the model may capture noise present in the training data, leading to bad performance on unseen examples.

The first convolutional layer of this deep architecture has, concretely, the input dimensions (225, 225, 3), for width, height, and channels, and it contains 32 filters. Each filter size is (3, 3). ReLU is the activation function. For the first max pooling layer, the pool size is (2, 2) while the output dimensions are (112, 112, 32) (see Fig. 5). The second convolutional layer has 64 filters with a size (3, 3) while the activation function is ReLU, for an output of (110, 110, 64). The second maximum pool layer, with a pool size (2, 2), has output dimensions (55, 55, 64). Again, a convolutional layer is used, with 128 filters, and size (3, 3). The applied activation function is ReLU. The output dimensions are (53, 53, 128). Another max pooling layer is added to the third convolutional layer. This one also has a pooling size of (2, 2), but output dimensions of 26×26. The next layer is the flattened one, with the output dimensions of (86528,) (see (4)). The eighth layer of this deep learning model is the fully connected one (the dense). It has 128 neurons, the activation function ReLU, and the main operation:

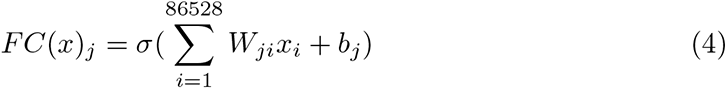

where *FC*(*x*)*_j_* represents the *j*-th element of the output vector produced by the fully connected layer when given the input vector *x*. The notation *σ* denotes the activation function of the fully connected layer core. After that, instructions represent the weighted sum of the input features, *x_i_*, multiplied by their corresponding weights *W_ji_*. The summation runs over all *i* features, and the weights *W_ji_* represent the connection strengths between the *i*-th input feature and the *j*-th output neuron. The *b_j_* term is the bias associated with the *j*-th output neuron. It is added to the weighted sum before applying the activation function. The bias allows the model to learn an offset that helps in better fitting the data. The dropout layer, with a 0.5 rate, helps prevent overfitting by assessing a fraction of input units to 0 per update during the training. The fully connected layer contains 2 neurons, assuming it’s a classification task with 2 classes, and uses the softmax activation function [58] (see (5)). The main operation is:

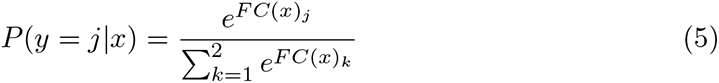

where *P* represents the probability that the input *x* belongs to class *j*. In the context of softmax activation, the output of the fully connected layer is transformed into a probability distribution over the classes. The *e^FC^*^(*x*)*j*^ term is the exponential of the *j*-th element of the output vector from the fully connected layer. The represented sum normalizes the probabilities, ensuring that the sum of probabilities across all classes equals 1. This normalization step is crucial to interpret the output as probabilities. Finally, considering the compilation of the model, optimizer Adam is used, with a learning rate of 0.001 and the loss function binary cross-entropy. The accuracy metric is considered essential here. The summary of the model provides information about the architecture, including the number of parameters in each layer. It is important to note that the dropout layer introduces randomness during training, and the specific weights and biases are learned through the entire training process via backpropagation.

Compared to other models, this algorithm has several distinct advantages. The Bionnica model is simple in terms of implementation and comprises only a few essential layers. In contrast, other neural network architectures like AlexNet or ResNet-50 are more complex. Despite having only a few layers, Bionnica delivers high-performance results and trains the data more quickly. It is a highly customizable model, allowing the addition and removal of layers, the adjustment of hyperparameters, and the exploration of different configurations suitable for digital biopsy processes. Bionnica is more efficient than architectures like ResNet-50 or AlexNet in terms of computation time (see Fig. 6), making it compatible with limited computational resources. The use of dropout regularization helps prevent overfitting.

**Fig. 6.**
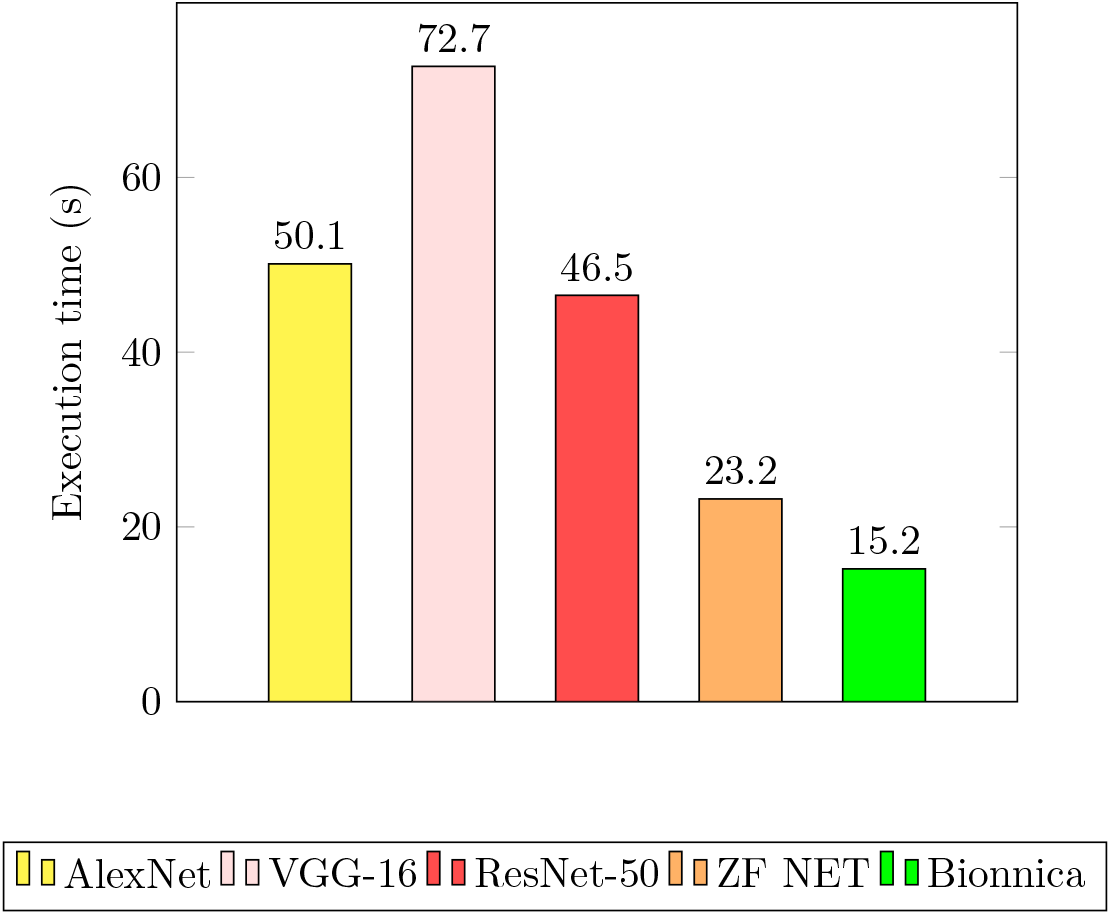
Execution times for models’ construction, training stage, and evaluation metrics’ calculation for all deep learning architectures.

However, there are some disadvantages, primarily related to the model’s depth. Relatively shallow neural networks cannot capture hierarchical features so well but require less computational power. Performance might be limited if the model’s simplicity is not well-structured. The proposed model has pre-trained models available, which means it doesn’t need to be fine-tuned for new polyps’ observations. Pre-trained models can save time and data for training, but they do not guarantee compatibility with new datasets.

## 4 Results and Discussion

The implementation details related to the results section are intended to evaluate the models developed using performance metrics such as ROC-AUC, sensitivity, precision, and F1-score. The execution time of the models is also measured.

In the evaluation of these neural network models for polyp classification, several key observations emerge. First, Bionnica and ZF NET exhibit the most efficient execution times among the models (Fig. 6), reflecting their ability to process data quickly. On the other hand, VGG-16 records the highest execution time, which could pose challenges for real-time applications.

Moving on to the metrics (Table 3), sensitivity, F1-score, and precision are the most popular and are used to assess autonomous polyp classifiers. In the context of polyps, a high ROC-AUC score means that the model is better at assisting clinicians in making informed decisions about the likelihood of malignancy. This can lead to more accurate diagnoses, potentially reducing unnecessary steps and improving patient outcomes [59].

**Table 3.** Performance evaluation metrics’ values for classifying colorectal polyps into premalignant and benign.

|  | **ZF NET** | **VGG-16** | **AlexNet** | **ResNet-50** | **Bionnica** |
| --- | --- | --- | --- | --- | --- |
| **ROC-AUC** | 74.1% | 72.7% | 70.3% | 69.9% | 85.5% |
| **F1-Score** | 84.2% | 80.8% | 77.4% | 84.2% | 86.1% |
| **Precision** | 79.0% | 74.7% | 70.7% | 77.1% | 80.0% |
| **Sensitivity** | 90.1% | 88.1% | 85.5% | 92.8% | 93.2% |

The deep architectures Bionnica and ResNet-50 deliver the highest sensitivity scores (above 92% for both models), indicating their ability to correctly identify a substantial proportion of premalignant polyps. Deep architectures, VGG-16 and ZF NET, also achieve commendable sensitivity figures (above 88% in both models), though slightly lower than the leading models. Bionnica and ZF NET achieve the highest precision scores, highlighting their effectiveness in minimizing false positives when categorizing polyps as malignant. In contrast, VGG-16 and ResNet-50 exhibit somewhat lower precision, suggesting a relatively higher rate of false-positive premalignant polyp cases.

In terms of ROC-AUC, Bionnica excels with the highest scores (85.5%), signifying its strong discriminative ability between premalignant and benign polyps (see Fig. 7). ZF NET, VGG-16, and AlexNet also produce respectable ROC-AUC values (around 70% are the areas), albeit below the top-performing model.

**Fig. 7.**
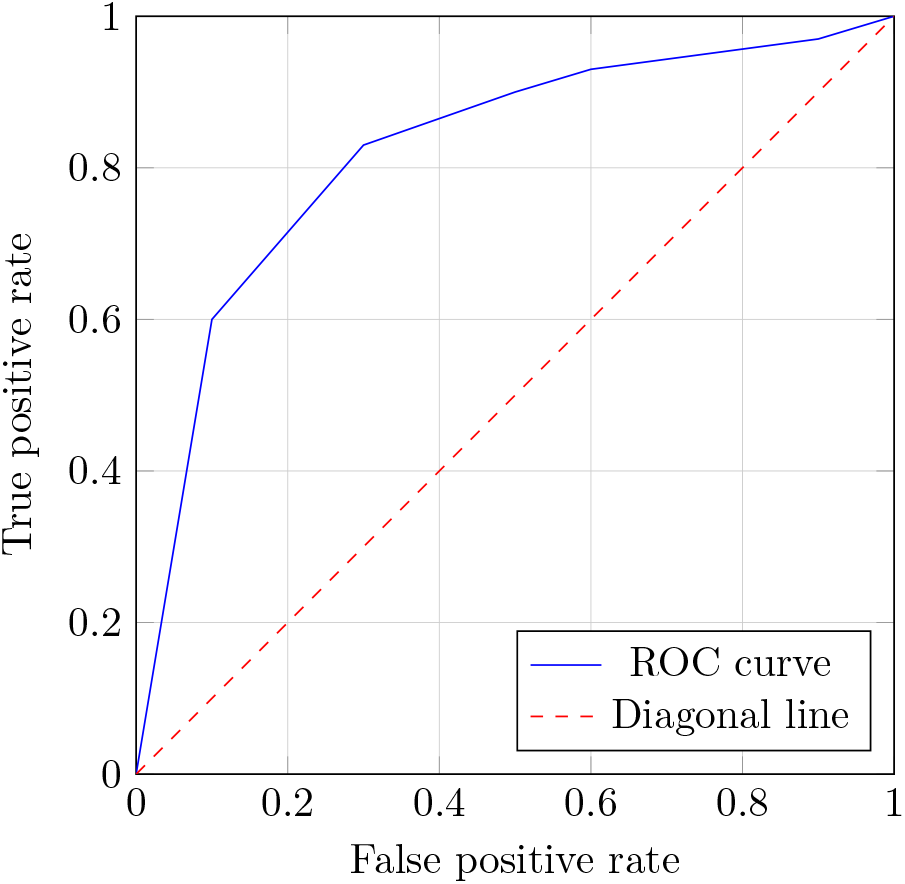
For rule-based enhanced Bionnica, the area under the ROC curve is 85.5%.

A K-fold cross-validation is a procedure that evaluates the performance of neural network models’ classification. The technique implies portioning the dataset into *k* folds that are around the same size [60]. There are *k* = 15 repetitions of the training and assessment process in our experiment. The model is tested on the remaining 1 fold after being trained on k-1 folds in each iteration. The performance metric, accuracy, is calculated for each iteration, resulting in *k* different values.

Bionnica has the highest accuracy values for almost every data section (see Fig. 8). ZF NET provides the best result for the sixth portion (above 78%) and the eleventh portion (almost 80%). AlexNet deep learning architecture comes after, with all the accuracies above 72%.

**Fig. 8.**
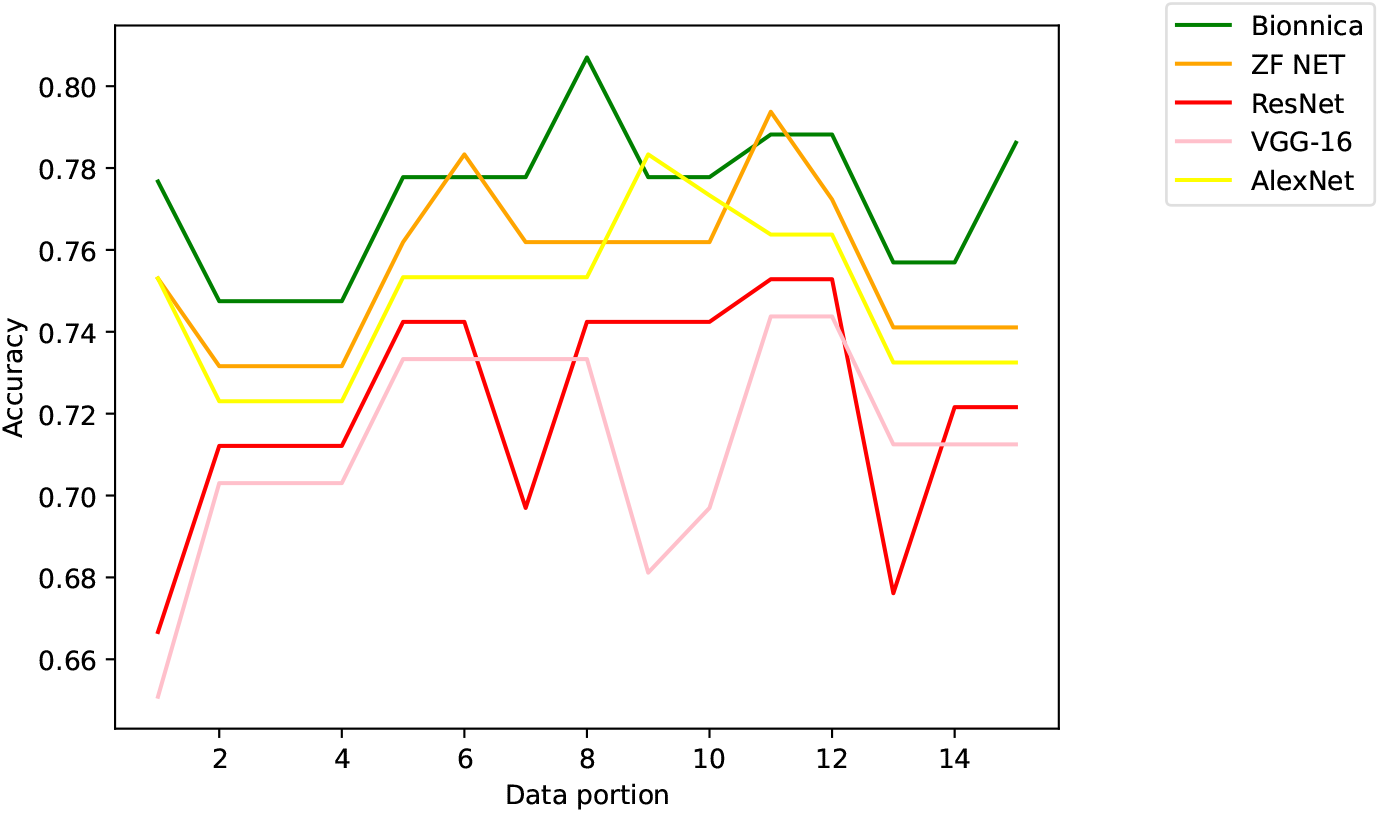
Accuracy values for each neural network architecture’s partition of k-fold cross-validation.

The Bionnica deep model generalizes well to different subsets of the data. This is a positive sign, indicating that the model is not overly sensitive to the specific data used for training and testing. Only robust models tend to perform well across various scenarios and data distributions, making them reliable in real-world applications [61]. High accuracy suggests that the models can reliably distinguish between benign and premalignant colorectal polyps.

## 5 Conclusions

In this paper, we built a custom-based deep-learning architecture for classifying colorectal polyps into premalignant and benign. We analyzed multiple neural network models for the premalignant and benign polyps classification for a publicly available dataset with patients, focusing on performance evaluation metrics and models’ execution times, to improve the traditional methods. The model, integrating collaborative filtering within a medical context, benefits from inferred user preferences alongside image features, enhancing its ability to make specific classifications for colorectal image analysis. Collaborative filtering enables the model to adapt to individual user behavior, improving the accuracy and relevance of its outputs in medical scenarios.

The results have illuminated several strong points, as well as areas that warrant improvement and potential avenues for enhancement. Among the strong points, ZF NET and Bionnica stood out for their efficient execution time, making them well-suited for real-time applications where prompt results are crucial. Additionally, high sensitivity scores were observed in Bionnica, ZF NET, and ResNet-50, signifying their ability to correctly identify a substantial percentage of premalignant polyps, which is essential in medical diagnosis. The ROC-AUC is especially relevant because it provides a comprehensive evaluation of a model’s ability to discriminate between different classes or conditions, which is crucial in distinguishing benign and malignant polyps during colonoscopy. The ZF NET achieved the second-highest F1-score, while Bionnica exhibited an excellent F1-score, the highest one, demonstrating their ability to strike a harmonious balance between precision and sensitivity, which is vital for accurate classification.

Fine-tuning the models to reduce false positives could enhance their diagnostic accuracy. Bionnica, enhanced with a rule-based layer, improved the performance metrics’ values, efficiency, and clinical applicability in the vital task of diagnosing premalignant polyps, ultimately improving patient care and healthcare concerns.

## Declarations

### Data Availability

The datasets analyzed during the current study can be accessed via the following link: https://www.depeca.uah.es/colonoscopy_dataset/. The dataset was made publicly available by the authors of [10]. We asked the authors about using the images. The images were captured from the white light frame and NBI videos. The listed images were used by us to perform further bioimage processing.

## Data Availability

All data produced in the present work are contained in the paper

